## Supplementary material for "Anxiety and Depression among Medical Doctors in Catalonia, Italy, and the UK during the COVID-19 Pandemic": S1 Appendix

This supplemental material has been provided by the authors to give readers additional information about their work.

**This document contains:**

Additional Tables ..... S2
- Table S1: Survey details following AAPOR survey disclosure guidelines ..... S2
- Table S2: Place of work among UK respondents ..... S3
- Table S3: Response rates by institutions ..... S4
- Table S4: Definition of key variables ..... S5
- Table S5. Original and final sample sizes ..... S6
- Table S6: Logit odds-ratios and 95% confidence intervals ..... S7

Additional Figures ..... S8
- Figure S1: Example email invitation June 2020 (RCPSG) ..... S8
- Figure S2: Example email invitation November 2020 (RCSEd) ..... S9
- Figure S3: Prevalence of anxiety and depression symptoms by intensity ..... S10

### Additional Tables

Table S1: Survey details following AAPOR survey disclosure guidelines

| BASIC DISCLOSURE ELEMENTS | DETAILS |
|---|---|
| Survey sponsor | Surveys were distributed online by the medical organizations (COMB, COMG, ANAAO-ASSOMED, FIMMG, RCSED, RCPSG) via their mailing lists. |
| Survey/Data collection supplier | Researchers from the universities of Cambridge, Exeter and Glasgow designed the survey in Qualtrics. |
| Population represented | Healthcare workers (medical doctors) in Catalonia, Italy and the UK. |
| Sample size | 5,275 |
| Mode of data collection | Online |
| Type of sample (probability/non-probability) | Non-probability (ANAAO-ASSOMED, FIMMG, RCSED, RCPS) and probability (COMB, COMG). |
| Start and end dates of data collection | May 29, 2020 to June 30, 2020.<br>November 1, 2020 to December 31, 2020. |
| Margin of sampling error for total sample | NA |
| Margin of sampling error for key subgroups | NA |
| Are the data weighted? | The data are not weighted. Weights are not suitable in our context because the available weights reflect the composition of the respective institutions regardless of the characteristics of their members, while our sample focuses on a subsample of the underlying population (see Table S5). |
| Contact for more information | Climent Quintana-Domeque, PhD<br>University of Exeter, Business School, Department of Economics<br> |

Note: More information on the data and replication files is available at https://sites.google.com/site/climentquintanadomeque/healthcare-workers-survey.

Table S2: Place of work among UK respondents

| | RCPSG Round 1 | RCPSG Round 2 | RCSEd Round 1 | RCSEd Round 2 |
|---|---|---|---|---|
| England | 35.5% | 41.4% | 74.7% | 71% |
| Northern Ireland | 2.2% | 4.5% | 4.3% | 7.1% |
| Scotland | 61.5% | 52.2% | 16% | 18.6% |
| Wales | 0.8% | 1.9% | 5% | 3.3% |

Round 1 corresponds to June 2020; round 2 corresponds to November 2020.

#### Table S3: Response rates by institutions

| Institution | June 2020 | November/December 2020 |
|---|---|---|
| COMB (Catalonia) | 706/5,062[a] | 688/5,062[a] |
| COMG (Catalonia) | 170/3,120 | 285/3,120 |
| Anaao-Assomed (Italy) | 862/23,379[b] | 614/23,379[b] |
| FIMMG (Italy) | 775/17,687 | 323/17,687 |
| RCPSG (Scotland) | 231/3,990 | 157/4,300 |
| RCSEd (Scotland) | 281/4,992 | 183/4,912 |

Note: COMB (Barcelona Medical Council) and COMG (Girona Medical Council) are medical councils; Anaao-Assomed (Union of physicians and healthcare executives) and FIMMG (Union of general practitioners) are medical unions; RCSEd (Royal College of Surgeons of Edinburgh) and RCPSG (Royal College of Physicians and Surgeons of Glasgow) are private medical associations.

[a] Of the 36,339 COMB members, we focused on 25,425 members who were under 70 years, available to be contacted via e-mail, and willing to be contacted. Within this group, COMB invited 5,062 members in June and November 2020 (19.9%).

[b] This includes a large number of members who are not medical doctors.

Note that computing the response rate is problematic for several reasons, including the following: (1) membership changes over time (especially in private associations); (2) in Anaao-Assomed and UK, members are not just medical doctors.

Table S4: Definition of key variables

| Variable | Definition |
|---|---|
| **Key outcome variables** | |
| Anxiety | =1 if Generalized Anxiety Disorder Assessment (GAD-7) ≥ 10, =0 otherwise |
| Depression | =1 if Patient Health Questionnaire-9 (PHQ-9) ≥10, =0 otherwise |
| **Demographic and survey information** | |
| Sex | =1 if woman; =0 if man |
| Age ≥ 60 | = 1 if age 60 or above; = 0 otherwise |
| Survey Round | = 1 if surveyed in November/December 2020; = 0 if surveyed in June 2020 |
| **Perceptions of workplace safety** | |
| Do not have necessary PPE | =1 if do not agree with the statement "My workplace is providing me with the necessary Protective Personal Equipment"; = 0 otherwise |
| Little concern | = 1 if strongly agree or somewhat agree to the statement "My workplace has shown little concern for my safety"; = 0 otherwise |
| Feeling vulnerable/exposed | = 1 if strongly agree or somewhat agree to the statement "I feel vulnerable and exposed at work"; = 0 otherwise |
| **Exposure to COVID-19** | |
| COVID-19 symptoms | = 1 if had COVID-19 symptoms; 0 if no COVID-19 symptoms |
| Directly treat COVID-19 patients | = 1 if respondent replies "yes" to the question "In the last week, did you directly look after COVID-19 patients?"; = 0 if respondent replies "no". |
| Help with COVID-19 related tasks | = 1 if respondent replies "yes" to the question "In the last week, have you been asked to help out with work related COVID-19 patients without treating them directly?"; = 0 if respondent replies "no". |
| COVID-19 related deaths in workplace | = 1 if respondents says that there are positive number of doctor, nurse, or other personnel deaths in response to the question "Are you aware of any COVID-19 deaths among healthcare workers in your workplace? "; = 0 otherwise. |
| **Health, health behaviors, and lifestyle** | |
| Poor health | =1 if respondent rates their general health as "fair", "bad", or "very bad"; = 0 if respondent rates their general health as "good" or "very good" |
| Underlying health conditions | = 1 if respondent has underlying health conditions; = 0 otherwise |
| Worked ≥ 40 hours | = 1 if respondent worked 40 hours or more in the past week; = 0 otherwise |
| Smokes | = 1 if respondent smokes; = 0 otherwise |
| Had vaccine this season | = 1 if had flu vaccine this season; = 0 otherwise |
| **Household composition** | |
| Lives with child under 5 | = 1 if lives with a child under 5; = 0 otherwise |
| Lives with someone over 60 | = 1 if lives with someone 60 or above; = 0 otherwise |
| **Other indicators** | |
| Occupational indicators | 7 occupations indicators (e.g. in the UK: Consultant, SAS doctor, Specialty registrar, Junior doctor core training, Junior doctor foundation year, General practitioner, General practitioner trainee), that = 1 if respondent's occupation equals that category; = 0 otherwise. |
| Institutional indicators | 6 institutional indicators (COMB, COMG, Anaao-Assomed, FIMMG, RCPSG, RCSEd) that = 1 if respondent belongs to that institution; = 0 otherwise. |

#### Table S5. Original and final sample sizes

| | Catalonia | | Italy | | UK | |
|---|---|---|---|---|---|---|
| | COMB | COMG | Anaao-Assomed | FIMMG | RCPSG | RCSEd |
| **Round 1** | | | | | | |
| Period | June 2020 | June 2020 | June 2020 | June 2020 | June 2020 | June 2020 |
| N (initial)[a] | 1,067 | 275 | 1,524 | 1,136 | 399 | 575 |
| N (excluding missing info)[b] | 886 | 213 | 1,223 | 858 | 333 | 461 |
| N (excluding duplicates)[c] | 866 | 207 | 1,190 | 840 | 328 | 451 |
| N (excluding different region)[d] | 828 | 195 | 1,121 | 794 | 261 | 321 |
| N (excluding other occupation)[e] | 742 | 174 | 873 | 780 | 245 | 292 |
| N (excluding other cases)[f] | 706 | 170 | 862 | 775 | 231 | 281 |
| **Round 2** | | | | | | |
| Period | Nov 2020 | Nov 2020 | Dec 2020 | Dec 2020 | Nov 2020 | Nov 2020 |
| N (initial)[a] | 1,023 | 432 | 1,021 | 460 | 228 | 404 |
| N (excluding missing info)[b] | 835 | 356 | 856 | 360 | 175 | 238 |
| N (excluding duplicates)[c] | 824 | 346 | 842 | 347 | 174 | 237 |
| N (excluding different region)[d] | 781 | 324 | 802 | 328 | 162 | 217 |
| N (excluding other occupation)[e] | 704 | 299 | 632 | 327 | 160 | 189 |
| N (excluding other cases)[f] | 688 | 285 | 614 | 323 | 157 | 183 |

[a] Raw data.

[b] Excluding missing information on sex, age, household composition, occupation and specialty.

[c] Excluding duplicates. Duplicates are identified as observations with the same IP address, sex, age, number of children below 5 years in the household, number of children 6-17 years in the household, number of adults 18-59 years in the household, number of adults 60 years and above in the household, occupation and specialty.

[d] Excluding respondents working in a different region/country (e.g. respondents in COMB working outside Catalonia, respondents in RCSEd working outside the UK).

[e] Excluding other occupations (e.g. biologists).

[f] Excluding other cases (e.g. retired, on leave, shielding).

#### Table S6: Logit odds-ratios and 95% confidence intervals

| | All | | Catalonia | | Italy | | UK | |
|---|---|---|---|---|---|---|---|---|
| | Anxiety | Depression | Anxiety | Depression | Anxiety | Depression | Anxiety | Depression |
| **Demographics** | | | | | | | | |
| Woman | 1.77*** | 1.76*** | 1.51* | 1.45* | 1.92*** | 2.00*** | 1.63* | 1.81** |
| | [1.50,2.07] | [1.49,2.09] | [1.09,2.11] | [1.05,2.01] | [1.56,2.36] | [1.59,2.52] | [1.05,2.52] | [1.18,2.77] |
| Age<60 | 1.49*** | 1.58*** | 2.19*** | 2.16*** | 1.29* | 1.25 | 2.27* | 3.96** |
| | [1.22,1.82] | [1.28,1.96] | [1.40,3.43] | [1.39,3.35] | [1.02,1.65] | [0.96,1.64] | [1.06,4.88] | [1.68,9.30] |
| Round 2 | 1.01 | 0.9 | 0.78 | 0.84 | 1.04 | 0.81 | 1.51 | 1.43 |
| | [0.86,1.18] | [0.76,1.06] | [0.59,1.04] | [0.63,1.10] | [0.83,1.30] | [0.63,1.05] | [0.95,2.38] | [0.91,2.23] |
| **Perceptions of workplace safety** | | | | | | | | |
| No necessary PPE | 1.39*** | 1.27* | 1.43* | 1.23 | 1.37* | 1.26 | 1.15 | 1.42 |
| | [1.15,1.68] | [1.04,1.56] | [1.01,2.02] | [0.87,1.72] | [1.07,1.77] | [0.95,1.66] | [0.61,2.17] | [0.77,2.61] |
| Feel vulnerable/exposed | 1.68*** | 1.72*** | 1.45* | 1.72*** | 1.77*** | 1.61*** | 1.98** | 2.40*** |
| | [1.41,2.00] | [1.43,2.06] | [1.05,2.01] | [1.26,2.35] | [1.39,2.24] | [1.23,2.09] | [1.19,3.28] | [1.47,3.91] |
| Little concern for safety | 1.29* | 1.36** | 1.35 | 1.43 | 1.25 | 1.52** | 1.64 | 1.04 |
| | [1.06,1.57] | [1.11,1.67] | [0.92,1.98] | [0.98,2.08] | [0.97,1.60] | [1.15,2.00] | [0.88,3.06] | [0.55,1.96] |
| **Exposure to COVID-19** | | | | | | | | |
| Had COVID-19 symptoms | 1.16 | 1.54*** | 1.31 | 1.93*** | 1.1 | 1.32 | 0.96 | 1.43 |
| | [0.97,1.39] | [1.29,1.84] | [0.98,1.76] | [1.46,2.55] | [0.84,1.44] | [0.99,1.76] | [0.59,1.55] | [0.91,2.25] |
| Directly treat | 1.32** | 1.32** | 1.39* | 1.55** | 1.29* | 1.31* | 1.34 | 1.12 |
| COVID-19 patients | [1.12,1.56] | [1.11,1.57] | [1.01,1.90] | [1.14,2.10] | [1.03,1.61] | [1.03,1.68] | [0.83,2.15] | [0.70,1.78] |
| Help w/ COVID-19 | 1.20* | 1.20* | 1.21 | 0.97 | 1.28* | 1.51** | 0.98 | 1.03 |
| related tasks | [1.02,1.42] | [1.01,1.43] | [0.89,1.64] | [0.72,1.31] | [1.02,1.60] | [1.18,1.92] | [0.61,1.57] | [0.65,1.62] |
| ≥1 COVID-19 death | 1.20* | 1.15 | 1.23 | 1.36 | 1.17 | 1.09 | 1.29 | 1.1 |
| in workplace | [1.02,1.41] | [0.97,1.37] | [0.88,1.73] | [0.98,1.88] | [0.95,1.44] | [0.86,1.38] | [0.83,1.98] | [0.73,1.68] |
| **Health status and behaviors** | | | | | | | | |
| Poor health | 2.58*** | 3.35*** | 2.31*** | 2.94*** | 2.85*** | 3.92*** | 2.65** | 3.57*** |
| | [2.13,3.13] | [2.76,4.06] | [1.70,3.14] | [2.19,3.94] | [2.18,3.73] | [2.96,5.18] | [1.31,5.39] | [1.81,7.06] |
| Underlying | 0.99 | 1.17 | 0.89 | 0.95 | 0.99 | 1.26 | 1.04 | 1.24 |
| health condition | [0.83,1.17] | [0.98,1.38] | [0.65,1.22] | [0.70,1.29] | [0.80,1.24] | [0.99,1.60] | [0.63,1.72] | [0.77,2.00] |
| Worked ≥ 40 hours | 1.44*** | 1.27** | 1.32 | 1.25 | 1.55*** | 1.25 | 1.12 | 1.27 |
| | [1.21,1.70] | [1.07,1.52] | [0.95,1.82] | [0.92,1.70] | [1.25,1.93] | [0.99,1.59] | [0.66,1.90] | [0.76,2.13] |
| Smokes | 1.04 | 1.50*** | 0.8 | 1.19 | 1.08 | 1.58** | 2.06 | 2.44 |
| | [0.83,1.30] | [1.20,1.87] | [0.51,1.25] | [0.80,1.78] | [0.82,1.42] | [1.19,2.11] | [0.79,5.35] | [0.98,6.10] |
| Had flu vaccine | 0.98 | 1.16 | 1.03 | 0.99 | 0.87 | 1.27* | 1.56 | 1.37 |
| this season | [0.84,1.15] | [0.98,1.36] | [0.77,1.39] | [0.75,1.31] | [0.71,1.08] | [1.01,1.61] | [0.95,2.57] | [0.86,2.19] |
| **Household composition** | | | | | | | | |
| Lives w/ child u5 | 1.32* | 1.03 | 1.4 | 1.1 | 1.29 | 1.14 | 1.22 | 0.62 |
| | [1.06,1.63] | [0.82,1.29] | [0.96,2.06] | [0.75,1.62] | [0.96,1.72] | [0.83,1.58] | [0.65,2.29] | [0.31,1.24] |
| Lives w/ someone over 60 | 1.04 | 0.94 | 1.59* | 1.32 | 0.91 | 0.79 | 0.69 | 0.89 |
| | [0.86,1.25] | [0.77,1.15] | [1.09,2.33] | [0.91,1.92] | [0.73,1.15] | [0.61,1.02] | [0.28,1.69] | [0.39,2.01] |
| Mean dependent variable | 0.20 | 0.19 | 0.15 | 0.17 | 0.26 | 0.21 | 0.14 | 0.16 |
| Observations | 4,993 | 4,993 | 1,737 | 1,737 | 2,444 | 2,447 | 806 | 806 |

Notes: Anxiety=1 if Generalized Anxiety Disorder Assessment (GAD-7) ≥ 10, =0 otherwise; Depression=1 if Patient Health Questionnaire-9 (PHQ-9) ≥ 10, =0 otherwise. All models include occupational fixed effects and institutional fixed effects. * p<0.05 ** p<0.01 *** p<0.001. 95% confidence intervals in square brackets.

### Additional Figures

Figure S1: Example email invitation June 2020 (RCPSG)

Dear Dr [redacted],

***- Sent on behalf of Professor Jackie Taylor, President -***

The Royal College of Physicians and Surgeons of Glasgow is collaborating with a group of university researchers at the University of Glasgow, the University of Cambridge and the University of Exeter to understand how healthcare workers are being affected by the COVID-19 pandemic.

If you have 10 minutes to spare, please help us fill in the following short and anonymised survey by visiting the following link:

https://uebs.eu.qualtrics.com/jfe/form/SV_2ohMr8isdmO6AbX

More information about the survey and the research team conducting this survey can be found here:

https://sites.google.com/view/hcws

We thank you in advance for your contribution to this research project by participating in the survey.

Your anonymised responses will help improve our understanding of what measures should be taken to help healthcare workers adjust to the consequences of this crisis.

Many thanks,

David Thomson

David Thomson
**Global Engagement Officer, Membership and Global Engagement Unit**
**Royal College of Physicians and Surgeons of Glasgow**
232 - 242 St Vincent Street, Glasgow, G2 5RJ
T + 44 (0)141 221 6072 | F + 44 (0)141 221 1804

Figure S2: Example email invitation November 2020 (RCSEd)

THE ROYAL COLLEGE OF SURGEONS OF EDINBURGH

HCWS UNIVERSITY OF CAMBRIDGE UNIVERSITY OF EXETER University of Glasgow

**The Effects of a Second Wave on Healthcare Workers**

In June our Members were invited to participate in a study conducted by a group of researchers at the University of Glasgow, the University of Cambridge and the University of Exeter to understand how healthcare workers are being affected by the COVID-19 pandemic.

In order to compare the effects of the first and second wave, they would like to invite our Members to participate in a second survey. The survey will help understand what measures should be taken to help healthcare workers adjust to the consequences of this crisis.

If you have 10 minutes to spare, can you please fill in the following short and anonymised survey.

More information about the survey and the research team conducting this survey can be found here.

We thank you in advance for your contribution to this research project by participating in the survey.

START SURVEY

Figure S3: Prevalence of anxiety and depression symptoms by intensity

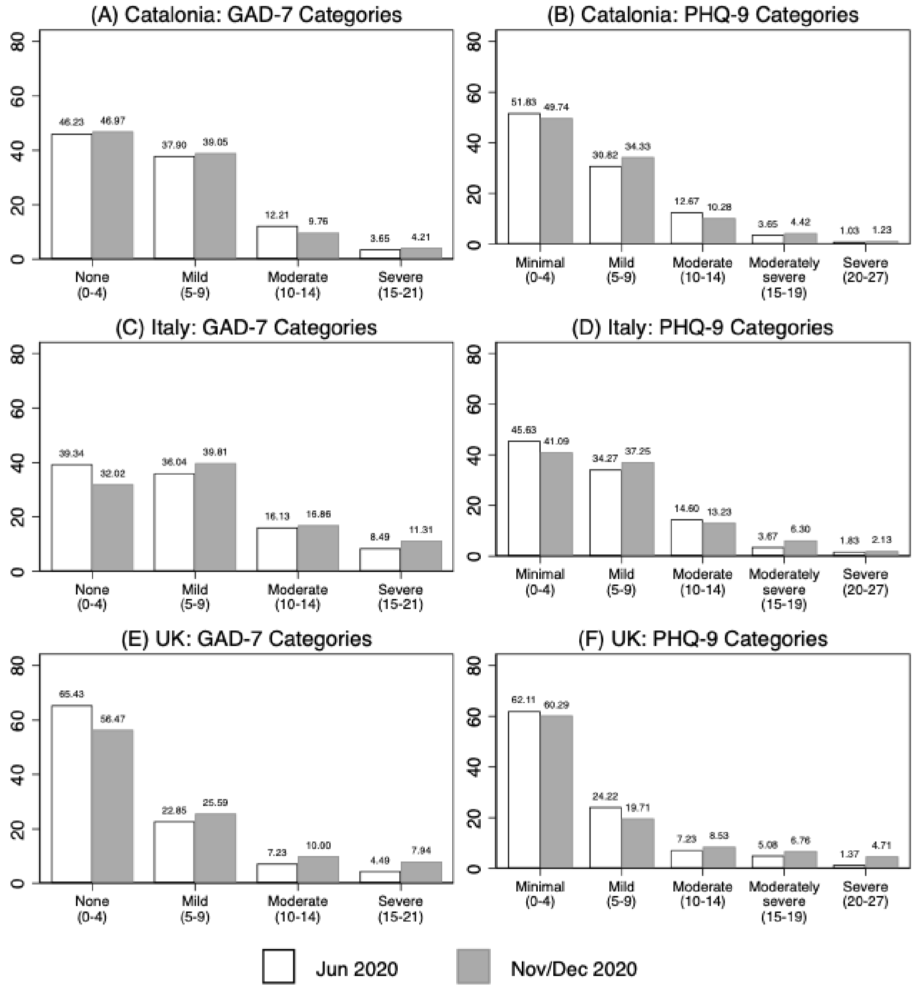
